# Estimating the probability of New Zealand regions being free from COVID-19 using a stochastic SEIR model

**DOI:** 10.1101/2020.04.20.20073304

**Authors:** Rafal Bogacz

**Affiliations:** MRC Brain Network Dynamics Unit, University of Oxford, UK

## Abstract

This report describes a method for estimating the probability that there are no infected or pre-symptomatic individuals in a populations on a basis of historical data describing the number of cases in consecutive days. The method involves fitting a stochastic version of Susceptible Exposed Infected Recovered model, and using the model to calculate the probability that the number of both exposed and infected individuals is equal to 0. The model is used to predict the current probabilities for all District Health Boards in New Zealand. These probabilities are highly correlated with the number of days with no new cases of COVID-19.

## Introduction

As the number of new cases of COVID-19 declines in New Zealand, the government is faced with a decision when to reduce the restrictions and release the country or its parts from lock-down. To assist in such a decision, it may be helpful to know the estimate for a probability of a particular region being free of COVID-19. This report presents a method for estimating such a probability on the basis of a simple model and presents results for individual District Health Boards (DHB) in New Zealand.

A simple model is employed which makes several simplifying assumptions. Although these assumptions may not be satisfied, they make the model mathematically tractable, and thus allows estimation of its parameters. In particular, the model makes the following assumptions.

- Since the Ministry of Health reports daily numbers of new cases in each DHB, the model treats each DHB as a separate population, thus assuming that individuals do not move between DHB (following government instructions not to travel).
- The model assumes that the behaviour of people is the same in the whole New Zealand and throughout the analysed period. Thus a single set of model parameters is fit to data from 25 March 2020 (when the lockdown was introduced in New Zealand) to 18 April 2020.
- Since the number of infected individuals in New Zealand is a small fraction of the entire population, the model assumes that the number of susceptible individuals is constant within the analysed period.
- The model assumes that infected individuals are eventually identified, and from this time, become fully isolated and no longer infect others.
- Since the incubation period of COVID-19 is around 5 days [1], the individuals who come in contact with the virus first enter a presymptomatic phase lasting on average 5 days to which we refer as an “exposed state”, and only then became infectious.

Given the above assumptions the results of the analysis need to be treated with caution. Nevertheless, it us hoped the the presented method may be also refined for more realistic set of assumptions.

## Methods

### Description of the model

We considered a stochastic version of Susceptible Exposed Infected Recovered (SEIR) model [2]. It assumes that each individual can be in one of 4 states, and move between as illustrated in Figure 1A. We denote by *E*_*t*_ and *I*_*t*_ the number of presymptomatic and infectious people on day *t*, and by Δ*E*_*t*_ and Δ*I*_*t*_ the number of people becoming presymptomatic or infectious. We do not have data on these variables, so we assume they are hidden to us and we will infer their values. On the other hand, we denote by Δ*R*_*t*_, the number of people being reported as a new case on day *t*, and we assume that this observed number corresponds to the amount of people moving to state *R* where they no longer infect others (as they become fully isolated). Thus in the presented model the cases reported correspond to individuals moving to state *R* rather than *I* as typically assumed, because we assume that the individuals are more likely to infect others when they are unaware that they have COVID-19, rather than after diagnosis when they become particularly careful. Indeed, James et al. recently wrote that the control measures that specifically target confirmed cases, such as isolation, “could be modelled by a larger reduction in transmission rates for confirmed cases” [3]. Here we for simplicity assume that this transmission rate is reduced to 0.

**Figure 1:**
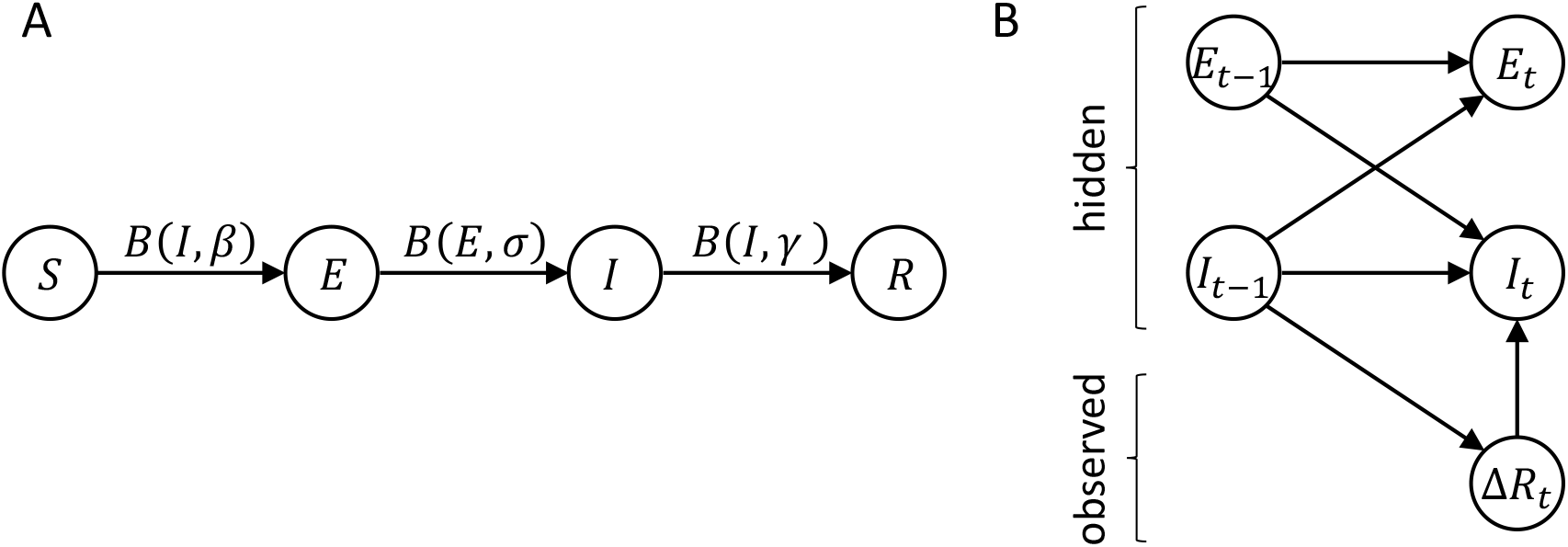
Model. A) Structure of SEIR model. Circles denote compartments, while arrows denote the movement of individuals between compartments, and the labels above arrows indicate the distributions of the number of people moving per day. B) Probabilistic model. Circles denote random variables, while arrows denote dependencies among them.

The changes in model variables over time are given by:

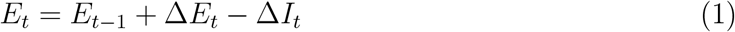

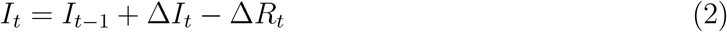

The model assumes that every day each infectious individual can infect another person with probability *β*. Hence the number of people becoming presymtomatic is a random variable with a binomial distribution *B*:

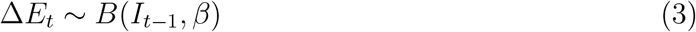

Additionally, we assume that presymptomatic and infectious people progress to the next state in Figure 1A (i.e. infectious and reported) with probabilities *σ* and *γ* respectively:

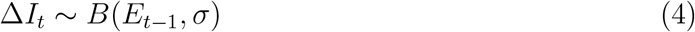

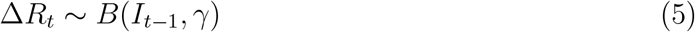

### Estimating the probability of being virus free

We wish to compute a probability distribution of variables *E*_*t*_ and *I*_*t*_ given data on the number of reported cases upto day *t*, which we denote by Δ*R*_1:*t*_. According to the definition of conditional probability:

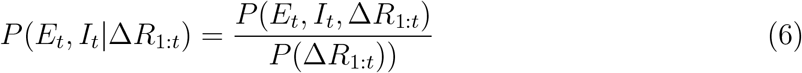

where

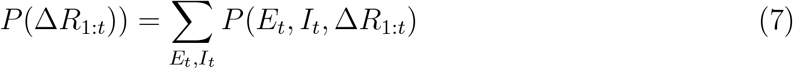

To compute *P* (*E*_*t*_, *I*_*t*_, Δ*R*_1:*t*_) we take advantage of the fact that the dependencies between variables in the model (Figure 1B) have a similar structure as in a hidden Markov model, so this probability can be computed recursively using an algorithm analogous to the forward algorithm. Thus this probability can be decomposed into a sum of probabilities of disjoint events:

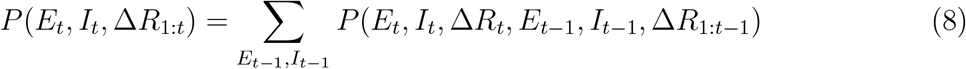

Using the definition of a conditional probability, the probability on the right hand side of the above equation can be written as a chain of conditional probabilities:

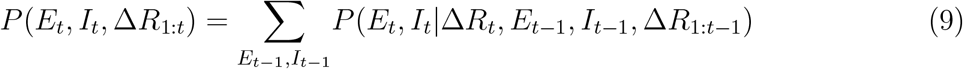

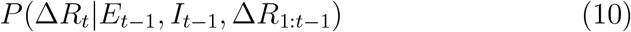

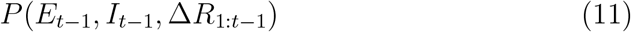

Noting that the variables depend only on subset of variables as illustrated in Figure 1B, the above expression can be simplified to:

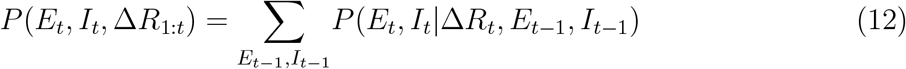

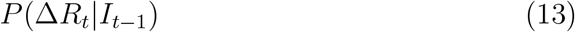

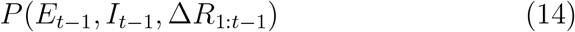

To compute the term on the right hand side of Equation 12, we note that *E*_*t*_ and *I*_*t*_ are determined by changes in these variables (Equations 1-2). To find how Δ*E*_*t*_ is related with other variables, we can add Equations 1 and 2, and we get:

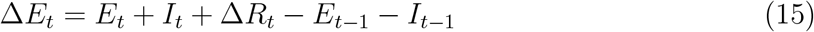

Using Equations 15, 2-5 we obtain:

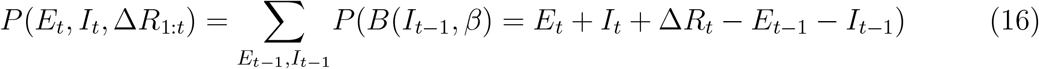

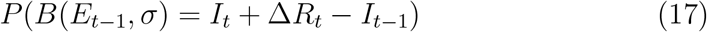

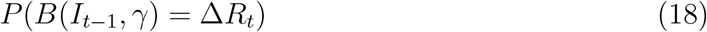

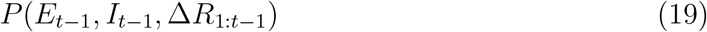

The above formula allows recursively computing *P* (*E*_*t*_, *I*_*t*_, Δ*R*_1:*t*_) from the corresponding probability on the previous day. In the presented analyses these probabilities were computed for the values of *E*_*t*_ and *I*_*t*_ from 0 to *n*, and in the first step we used uninformative flat prior 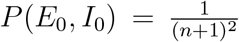 Once the joint probability *P* (*E*_*t*_, *I*_*t*_, Δ*R*_1:*t*_) is computed for the last available day, we compute the posterior probability from Equation 6, and report the probability of the population being virus free *P* (*E*_*t*_ = 0, *I*_*t*_ = 0|Δ*R*_1:*t*_).

### Estimating model parameters

To perform the calculation in the previous section, the values of parameters *β, σ* and *γ* need to be known. We set *σ* = 0.2 since the average incubation period of COVID-19 is around 5 days. We assume that *β* and *γ* are the same for the whole New Zealand. To estimate *β* and *γ*, we find the values which maximize the product of probabilities of observed data (Equation 7) across DHB. This maximum is found through a numerical optimization employing a simplex algorithm [4] with initial values *β* = *γ* = 0.3.

To assess accuracy of this parameter estimation procedure we tested if the correct values of parameters can be recovered from artificially generated data. We generated 20 artificial datasets, each containing a similar amount of data that is available on the current history of cases in New Zealand. For each dataset we randomly generated *γ* from uniform distribution between 0.1 and 0.4, and generated *β* from uniform distribution between 0.05 and *γ* + 0.05. With these parameters we generated 20 sequences (corresponding to 20 DHB) of Δ*R*_*t*_. For each DHB, the initial numbers of presymptomatic and infectious individuals *E*_0_ and *I*_0_ were generated from uniform distribution between 1 and 10. The model was then simulated for 21 days according to Equations 1-5. For each dataset we estimated *β* and *γ* and compared with true values from which data were generated.

Left and middle displays in Figure 2 compare the estimated values of parameters with the true values. There is a significant correlation between true and recovered values (for *β*: *r* = 0.85, *p* < 0.01, for *γ*: *r* = 0.86, *p* < 0.01). Nevertheless there is a bias in the estimation. The direction of this bias is not consistent, but depends on simulation parameters (the recovery presented in Figure 2 was done with *n* = 10, but for *n* = 20 the bias is in the opposite direction, not shown). Nevertheless the right display in Figure 2 shows that the difference between parameters is accurately estimated (*r* = 0.97, *p* < 0.01). It is not surprising that this difference is recovered more accurately, because it determines whether the number of cases increases or decreases over days, which is a salient feature in the data. By contrast, the individual values of *β* and *γ* influence less salient features (e.g. a probability of a case after a long period with no cases), so they are more difficult to estimate.

**Figure 2:**
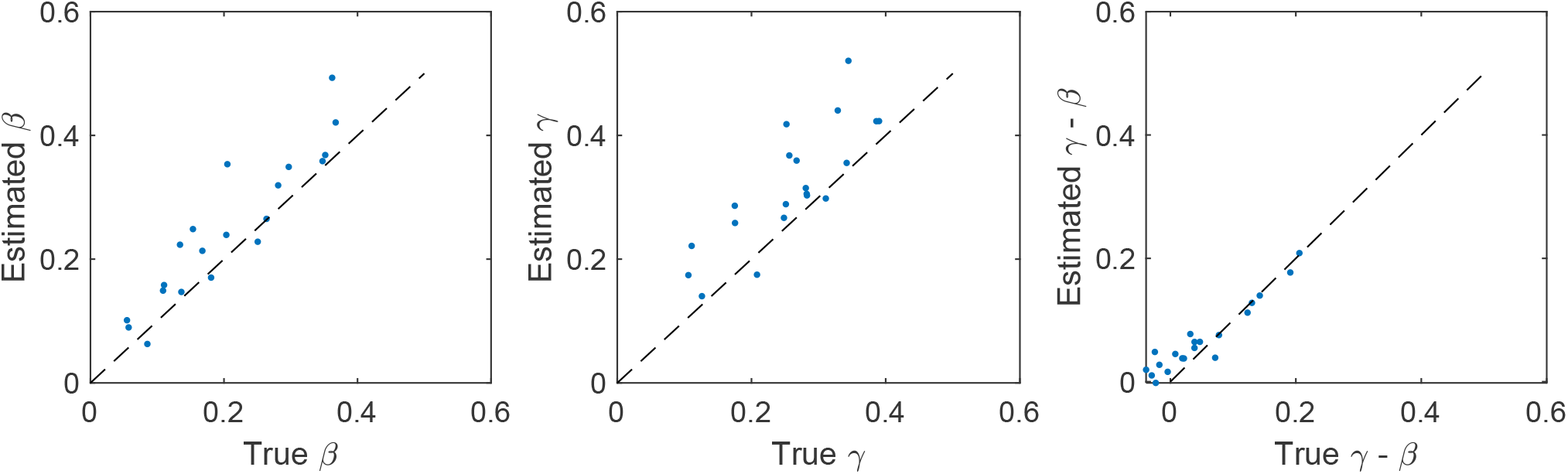
Comparison of true and estimated model parameters. Each dot corresponds to a single simulation, and the dashed lines indicate the identity line.

### Final estimation of probability

We estimated model parameters using the method described in the previous section with parameter *n* = 40 on the basis of data available at: https://www.health.govt.nz/our-work/diseases-and-conditions/covid-19-novel-coronavirus/covid-19-current-situation/covid-19-current-cases/. Form the available data we extracted the number of cases in each DHB on each day between 25 March and 18 April 2020.

Subsequently we used the fitted model to predict probability of the population being virus free *P* (*E*_*t*_ = 0, *I*_*t*_ = 0 Δ*R*_1:*t*_). To avoid the influence of the prior probability (that was set arbitrarily) on the estimate, for each DHB model was additionally run on the first week of data 10 times, i.e. the model was estimating the probability on the basis of extended sequence of new cases composed of: [Δ*R*_1:7_, Δ*R*_1:7_, Δ*R*_1:7_, Δ*R*_1:7_, Δ*R*_1:7_, Δ*R*_1:7_, Δ*R*_1:7_, Δ*R*_1:7_, Δ*R*_1:7_, Δ*R*_1:*t*_].

Due to uncertainty in estimates of individual parameters *β* and *γ* seen in Figure 2, we additionally estimated the probabilities of being virus free using a model with *β* = 0.1 and *β* = 0.4 and the parameter *γ* set on the basis of the estimated difference *γ* − *β*.

## Results

On the basis of data on the number of cases in New Zealand in the period from 25 March to 18 April 2020, we estimated parameters of the model as *β* = 0.26, *γ* = 0.42. The lower value of a rate with which individuals become infected *β* than the rate in which cases are reported *γ* corresponds to the decreasing number of cases over this period.

Table 1 gives estimated probability that the virus has been eradicated in individual DHBs (column 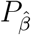). This probability is highly correlated with the number of days with no new cases, also given in Table 1 (*r* = 0.90, *p* < 0.01). This relationship is also illustrated in Figure 3. This relationship is not perfect, for example, the estimate for Whanganui is 44% despite only a single day with no cases, because this most recent case was preceded by 14 days with no cases in this DHB. In agreement with occurrence of cases after such long delays, even the regions without cases for 16 or 17 days have the probability estimate around 90% (rather than 100%).

**Table 1:** Estimates of the probabilities that different DHB in New Zealand are virus free. These estimates are given in column 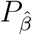.

| DHB | Number of cases | Days with no cases | $P_{\hat{\beta}}$ | $P_{\beta=0.1}$ | $P_{\beta=0.4}$ |
| --- | --- | --- | --- | --- | --- |
| Auckland | 150 | 0 | 0% | 0% | 0% |
| Bay of Plenty | 41 | 1 | 4% | 8% | 2% |
| Canterbury | 126 | 0 | 0% | 0% | 0% |
| Capital and Coast | 59 | 1 | 1% | 3% | 0% |
| Counties Manukau | 89 | 0 | 0% | 1% | 0% |
| Hawke's Bay | 38 | 6 | 15% | 21% | 11% |
| Hutt Valley | 15 | 13 | 74% | 77% | 71% |
| Lakes | 12 | 1 | 25% | 36% | 21% |
| Mid Central | 25 | 1 | 9% | 17% | 6% |
| Nelson Marlborough | 32 | 11 | 33% | 38% | 27% |
| Northland | 23 | 2 | 15% | 23% | 11% |
| South Canterbury | 13 | 1 | 17% | 27% | 14% |
| Southern | 194 | 1 | 0% | 0% | 0% |
| Tairāwhiti | 4 | 5 | 61% | 66% | 59% |
| Taranaki | 10 | 17 | 90% | 91% | 88% |
| Waikato | 167 | 0 | 0% | 0% | 0% |
| Wairarapa | 3 | 16 | 92% | 93% | 91% |
| Waitemata | 184 | 0 | 0% | 0% | 0% |
| West Coast | 4 | 13 | 91% | 92% | 90% |
| Whanganui | 8 | 1 | 44% | 54% | 38% |

**Figure 3:**
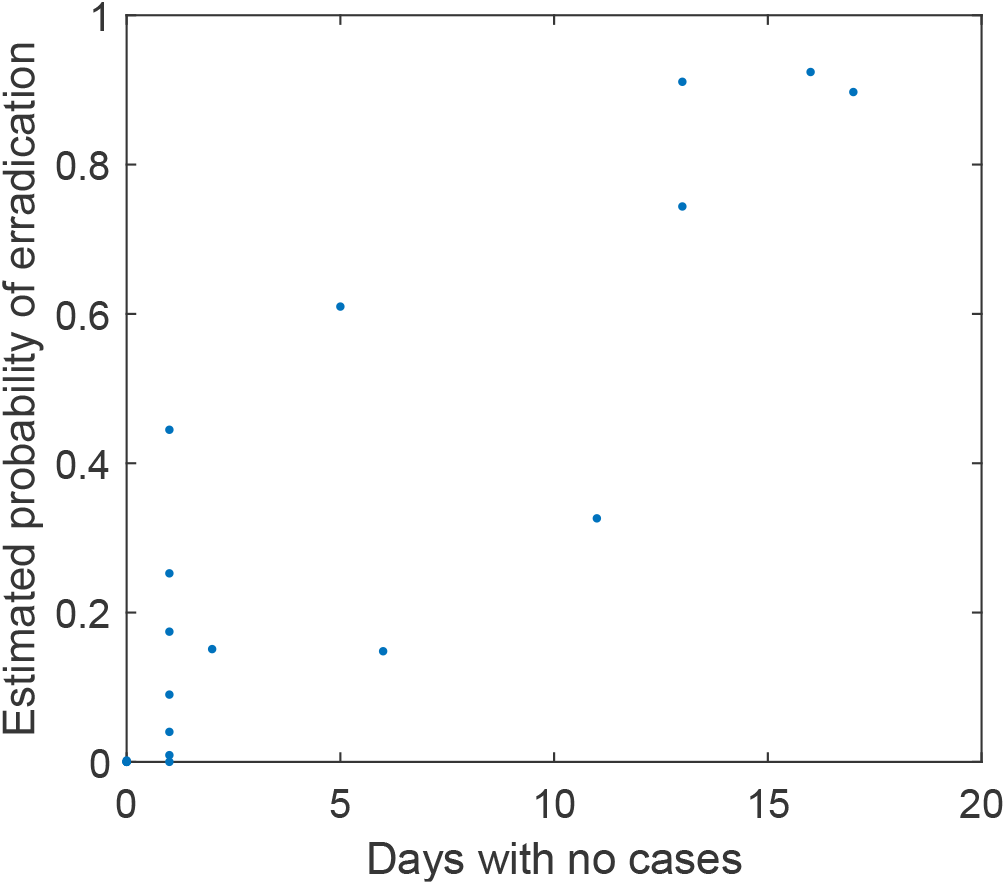
Relationship between the number of days with no new cases and the estimated probability of regions being virus free.

Two most right columns of Table 1 also list the probabilities computed with values of *β* differing form that estimated (and *γ* set to preserve the difference between estimated *γ* and *β*). These probabilities do not differ much, suggesting that the computation of these probabilities is robust to the biases in parameter estimates (Figure 2).

## Discussion

This report presented a method for estimating the probability of regions being virus free. This probability is highly correlated with the number of days without cases, suggesting that the number of case-free days is a useful quantity to consider while making decisions and informing the public. For example, Figure 4 illustrates how this quantity can be visualized.

The presented estimates have to be treated with great caution, because the model made many simplifying assumptions which are not satisfied in practice. First, the model assumes that all individuals with COVID-19 are symptomatic and identified, while another model of COVID-19 spread in New Zealand assumed probability 0.33 of individuals being subclinical [5]. It has been demonstrated that failing to detect accurately cases of disease increases the time one has to observe no new cases to be sure that a disease has been eradicated [6]. Nevertheless according to the website of the Ministry of Health (address in the Methods), the fraction of cases due to community transmission (4%) is much lower than the number of cases due to the contact with known individual (54%) which suggests that the majority of infectious individuals is being identified in New Zealand.

**Figure 4:**
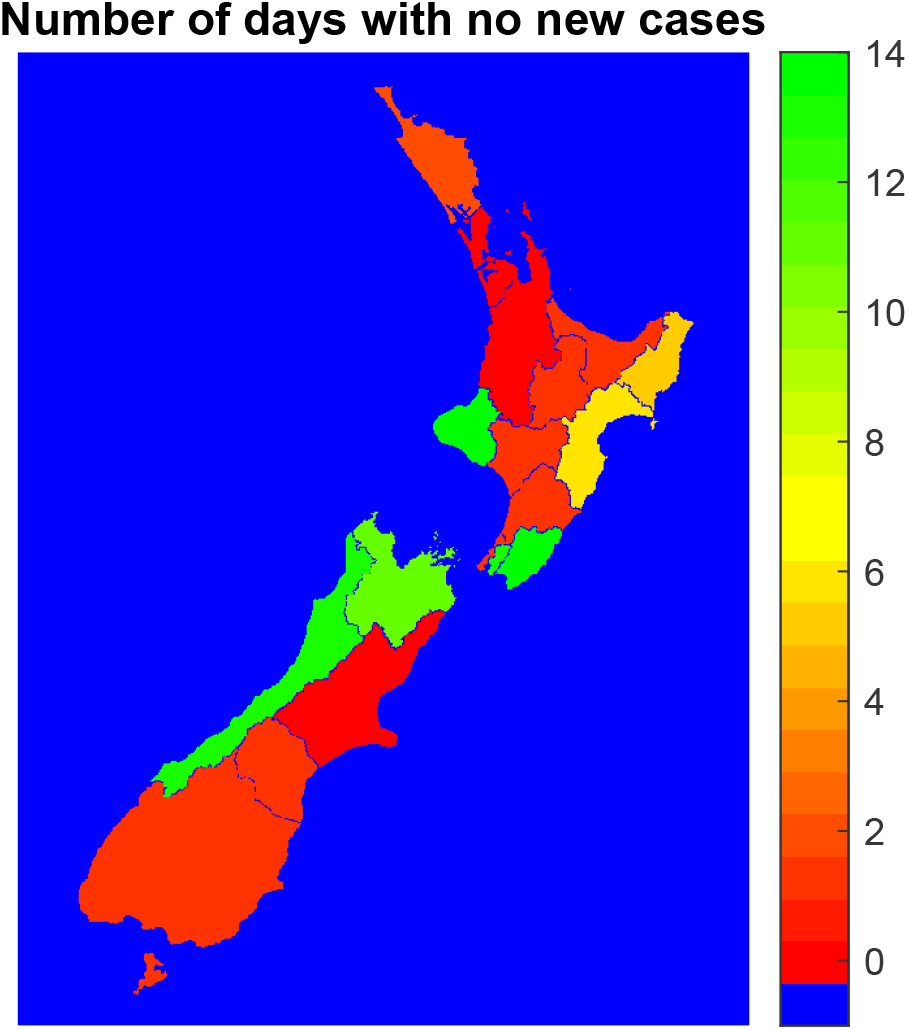
Map of New Zealand with different DHB colour coded according to the number of days with no new cases.

Second, the model assumed that once individuals become diagnosed, they are isolated, and no longer infect others. This assumption may also be not satisfied in practice. Third, the model assumes that there is no movement of individuals between DHB while some of the DHB (e.g. Auckland and Waitemata) include districts of the same city (e.g. Auckland), so the assumptions on travel of individuals between DHB could to be included to refine the model.

## Data Availability

The Matlab codes for estimating the probability of virus elimination, or for visualization of case-free days in New Zealand, are available from the author on request.

## Acknowledgement

The author thanks Medical Research Council UK for support.

